# Nosocomial Infections Among Patients with COVID-19, SARS and MERS: A Rapid Review and Meta-Analysis

**DOI:** 10.1101/2020.04.14.20065730

**Authors:** Qi Zhou, Yelei Gao, Xingmei Wang, Rui Liu, Peipei Du, Xiaoqing Wang, Xianzhuo Zhang, Shuya Lu, Zijun Wang, Qianling Shi, Weiguo Li, Yanfang Ma, Xufei Luo, Toshio Fukuoka, Hyeong Sik Ahn, Myeong Soo Lee, Enmei Liu, Yaolong Chen, Zhengxiu Luo, Kehu Yang, on behalf of COVID-19 evidence and recommendations working group

**Affiliations:** The First School of Clinical Medicine, Lanzhou University, Lanzhou 730000, China; Evidence-based Medicine Center, School of Basic Medical Sciences, Lanzhou University, Lanzhou 730000, China; Department of Respiratory Medicine, Children’s Hospital of Chongqing Medical University, Chongqing 400014, China; National Clinical Research Center for Child Health and Diseases, Ministry of Education Key Laboratory of Child Development and Disorders, China International Science and Technology Cooperation Base of Child Development and Critical Disorders, Children’s Hospital of Chongqing Medical University, Chongqing 400014, China; Chongqing Key Laboratory of Pediatrics, Chongqing 400014, China; Pediatric College, Chongqing Medical University, Chongqing 400014, China; School of Public Health, Chengdu Medical College, Chengdu 610500, China; Department of Pediatric, Sichuan Provincial People’s Hospital, University of Electronic Science and Technology of China, Chengdu 611731, China; Chinese Academy of Sciences Sichuan Translational Medicine Research Hospital, Chengdu 610072, China; School of Public Health, Lanzhou University, Lanzhou 730000, China; Emergency and Critical Care Center, the Department of General Medicine, Department of Research and Medical Education at Kurashiki Central Hospital, Japan; Advisory Committee in Cochrane Japan, Japan; Department of Preventive Medicine, Korea University College of Medicine, Seoul, Korea; Korea Cochrane Centre, Korea; Korea Institute of Oriental Medicine, Daejeon, Korea; University of Science and Technology, Daejeon, Korea; Lanzhou University, an Affiliate of the Cochrane China Network, Lanzhou 730000, China; Chinese GRADE Center, Lanzhou 730000, China; Key Laboratory of Evidence Based Medicine and Knowledge Translation of Gansu Province, Lanzhou University, Lanzhou 730000, China

**Keywords:** COVID-19, meta-analysis, nosocomial infection, rapid review

## Abstract

**Background:** COVID-19, a disease caused by SARS-CoV-2 coronavirus, has now spread to most countries and regions of the world. As patients potentially infected by SARS-CoV-2 need to visit hospitals, the incidence of nosocomial infection can be expected to be high. Therefore, a comprehensive and objective understanding of nosocomial infection is needed to guide the prevention and control of the epidemic.

**Methods:** We searched major international and Chinese databases Medicine, Web of science, Embase, Cochrane, CBM(China Biology Medicine disc), CNKI (China National Knowledge Infrastructure) and Wanfang database)) for case series or case reports on nosocomial infections of COVID-19, SARS(Severe Acute Respiratory Syndromes) and MERS(Middle East Respiratory Syndrome) from their inception to March 31st, 2020. We conducted a meta-analysis of the proportion of nosocomial infection patients in the diagnosed patients, occupational distribution of nosocomial infection medical staff and other indicators.

**Results:** We included 40 studies. Among the confirmed patients, the proportions of nosocomial infections were 44.0%, 36.0% and 56.0% for COVID-19, SARS and MERS, respectively. Of the confirmed patients, the medical staff and other hospital-acquired infections accounted for 33.0% and 2.0% of COVID-19 cases, 37.0% and 24.0% of SARS cases, and 19.0% and 36.0% of MERS cases, respectively. Nurses and doctors were the most affected among the infected medical staff. The mean numbers of secondary cases caused by one index patient were 29.3 and 6.3 for SARS and MERS, respectively.

**Conclusions:** The proportion of nosocomial infection in patients with COVID-19 was 44%. Patients attending hospitals should take personal protection. Medical staff should be awareness of the disease to protect themselves and the patients.

## Background

COVID-19 is a respiratory infectious disease caused by a novel coronavirus, SARS-CoV-2. The first batch of COVID-19 patients were found in China in December 2019(1). The disease is mainly transmitted through respiratory droplets and close contact, and all people are susceptible to it(2). SARS-CoV-2 is highly contagious(3), and has quickly spread to most countries and regions of the world. COVID-19 has become a global pandemic and has received great attention from all over the world(4,5). As of April 7, 2020, 1,214,466 confirmed cases of COVID-19 have been found in 211 countries and regions, causing 67,767 deaths(6).

The main clinical manifestations of COVID-19 are cough, fever and complications such as acute respiratory distress syndrome(1). Disease clusters and nosocomial infections have been reported(7,8). The proportion of nosocomial infections is high among diagnosed infections, and medical staff are at high risk of infection(8). One study on 44,672 patients showed that health workers accounted for 3.8% of the COVID-19 cases and five health workers died as a result of the infection(9). There is still no specific medicine for COVID-19, so preventing nosocomial infections is crucial.

This study compares the incidence of nosocomial infections during the COVID-19, SARS and MERS epidemics and analyzes the characteristics of the nosocomial infection, to enhance the understanding of nosocomial infection among medical and non-medical staff.

### Methods

### Search strategy

An experienced librarian searched the following databases from their inception to March 31, 2020 in the following electronic databases(10): the Cochrane library, MEDLINE (via PubMed), EMBASE, Web of Science, CBM (China Biology Medicine disc), CNKI (China National Knowledge Infrastructure), and Wanfang Data. We made no restrictions on language or publication status. We used the following search formula is as follow: (“Novel coronavirus” OR “2019-novel coronavirus” OR “Novel CoV” OR “2019-nCoV” OR “Wuhan-Cov” OR “2019-CoV” OR “Wuhan Coronavirus” OR “Wuhan seafood market pneumonia virus” OR “COVID-19” OR “SARS-CoV-2” OR “Middle East Respiratory Syndrome” OR “MERS” OR “MERS-CoV” OR “Severe Acute Respiratory Syndrome” OR “SARS” OR “SARS-CoV” OR “SARS-Related” OR “SARS-Associated”) AND (“Cross Infection” OR “Cross Infections” OR “Healthcare Associated Infections” OR “Healthcare Associated Infection” OR “Health Care Associated Infection” OR “Health Care Associated Infections” OR “Hospital Infection” OR “Nosocomial Infection” OR “Nosocomial Infections” OR “Hospital Infections” OR “hospital-related infection” OR “hospital-acquired infection”). We also searched clinical trial registry platforms (the World Health Organization Clinical Trials Registry Platform (http://www.who.int/ictrp/en/), US National Institutes of Health Trials Register (https://clinicaltrials.gov/)), Google Scholar (https://scholar.google.nl/), preprint platform (medRxiv (https://www.medrxiv.org/), bioRxiv (https://www.biorxiv.org/) and SSRN (https://www.ssrn.com/index.cfm/en/)) and reference lists of the included reviews to find unpublished or further potential studies. Finally, we contacted experts in the field to identify relevant trials. The search strategy was also reviewed by another information specialist. The details of the search strategy can be found in the **Supplementary Material 1**.

### Inclusion and exclusion criteria

We included case series studies and case reports about the proportion of cases of COVID-19, SARS and MERS who were infected in health facilities, about infections among medical staff and outbreaks in hospitals. Abstract, letter, new, guideline, articles for which we could not access all relevant data or full text were excluded.

### Study selection

After eliminating duplicates, two reviewers(Y Gao and X Wang) independently selected the relevant studies in two steps with the help of the EndNote software. Discrepancies were settled by discussion or consulting a third reviewer(Qi Zhou). In the first step, all titles and abstracts were screened using pre-defined criteria. In the second step, full-texts of the potentially eligible and unclear studies were reviewed to decide about final inclusion. All reasons for exclusion of ineligible studies were recorded. The process of study selection was documented using a PRISMA flow diagram (11).

### Data extraction

Two reviewers(R Liu and X Wang) extracted the data independently using a standardized data collection table. Any differences were resolved by consensus, and a third auditor checked the consistency and accuracy of the data. The following data were extracted: 1) basic information: title, first author, country, year of publication, and type of study; 2) population baseline characteristics: age and sex distribution, and sample size; and 3) the proportion of nosocomial infections, the proportion of patients with occupation of medical staff, and for studies on hospital outbreaks, the number of index cases and total infections.

### Risk of bias assessment

Two researchers (Z Wang and Q Shi) independently assessed the potential bias in each included study. The included studies were evaluated using appropriate assessment scales depending on the study type: for case control studies, the Newcastle-Ottawa Scale (NOS)(12), for cross-sectional studies and epidemiological surveys, the methodology evaluation tool recommended by the Agency for Healthcare Research and Quality (AHRQ)(13), and for case reports and case series, we used a methodology evaluation tool recommended by National Institute for Health and Care Excellence (NICE) (14).

### Data synthesis

We performed a meta-analysis of proportions for dichotomous outcomes (nosocomial infection among the confirmed cases, and infections among the health care workers), reporting the effect size (ES) with 95% confidence intervals (CI) by using random-effects models. Two-sided *P* values < 0.05 were considered statistically significant. Heterogeneity was defined as *P*<0.10 and *I*^*2*^>50%. All analyses were performed in STATA version 14.

### Quality of the evidence assessment

Two reviewers(Z Wang and Q Shi) assessed the quality of evidence independently using the Grading of Recommendations Assessment, Development and Evaluation (GRADE)(15-16). We produced a “Summary of Findings” table using the GRADEpro software. This table includes overall grading of evidence body for each prespecified outcome that is accounted in a meta-analysis. The overall quality can be downgraded for five considerations (study limitations, consistency of effect, imprecision, indirectness, and publication bias) and upgraded for three considerations (large magnitude of effect, dose-response relation and plausible confounders or biases). The overall quality of evidence will be classified as high, moderate, low or very low, which reflecting to what extent that we can be confident the effect estimates are correct.

As COVID-19 is a public health emergency of international concern and the situation is evolving rapidly, our study was not registered in order to speed up the process (17).

## Results

### Characteristics and quality of included studies

Our initial search revealed 2626 articles, of which 2598 were left after deleting the duplicates (*Figure 1*). After review the titles and abstracts, we screened the full texts of 66 articles, of which 40 were finally included (Table 1) (8,18-56). Four studies were about COVID-19, 25 studies about SARS, and 11 studies about MERS (*Table 1*). Sixteen studies described the number of nosocomial infections in a selected patient population, 16 studies described the situation of nosocomial infections among the staff of medical institutions, and 13 studies reported the number of nosocomial infections caused by one or more than one patient. The quality of included studies was very poor: all cross-sectional studies scored less than 8 out of 11 in the evaluation by the AHRQ tool, half case series studies scored less than 5 out of 8 in the evaluation by the NICE tool, and only one case-control study scored 6 by the NOS tool. The details of the risk of bias of included studies can be found in the **Supplementary Material 2**.

**Table 1.**
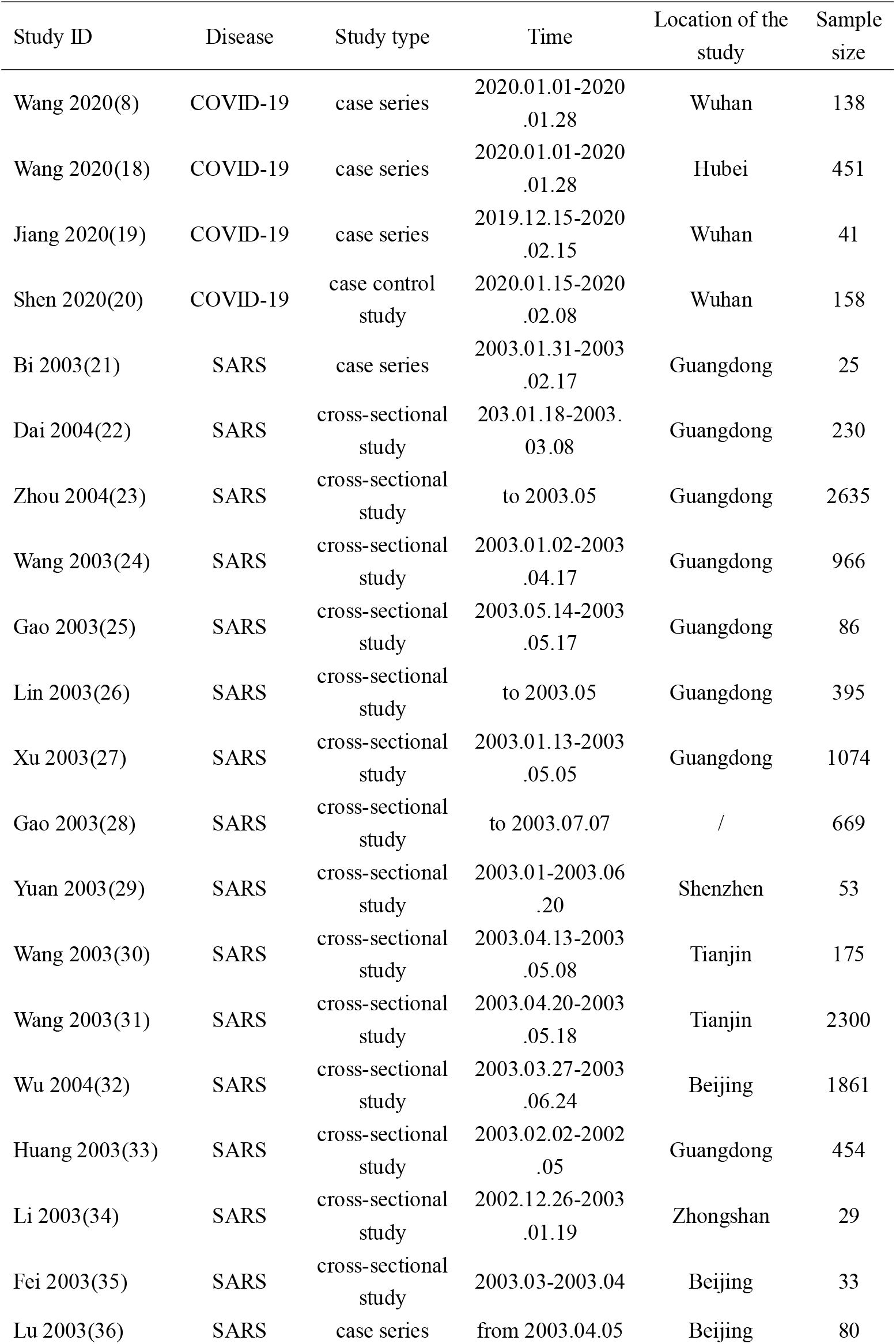
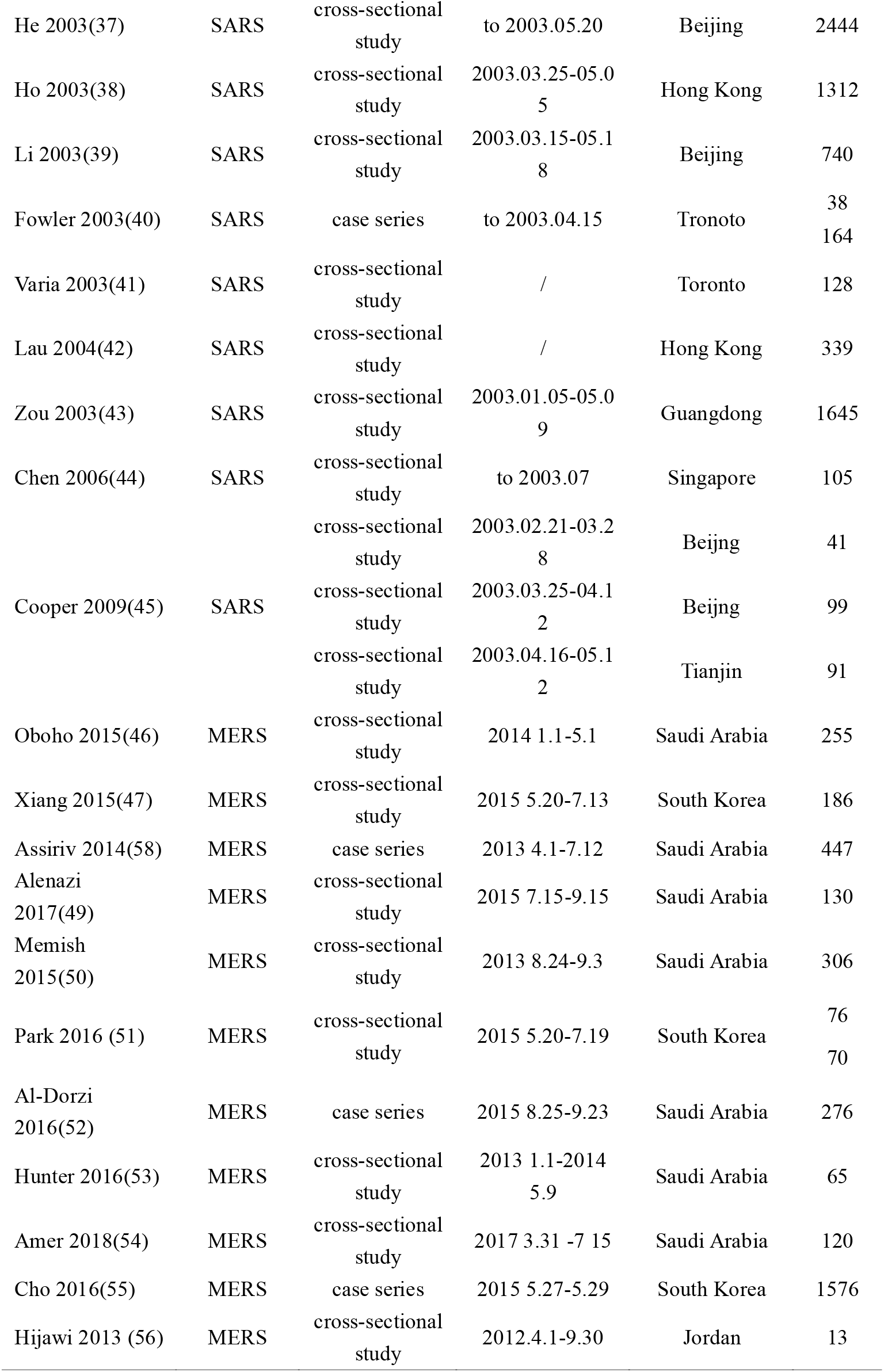
Characteristics of included studies

| Study ID | Disease | Study type | Time | Location of the study | Sample size |
| --- | --- | --- | --- | --- | --- |
| Wang 2020(8) | COVID-19 | case series | 2020.01.01-2020.01.28 | Wuhan | 138 |
| Wang 2020(18) | COVID-19 | case series | 2020.01.01-2020.01.28 | Hubei | 451 |
| Jiang 2020(19) | COVID-19 | case series | 2019.12.15-2020.02.15 | Wuhan | 41 |
| Shen 2020(20) | COVID-19 | case control study | 2020.01.15-2020.02.08 | Wuhan | 158 |
| Bi 2003(21) | SARS | case series | 2003.01.31-2003.02.17 | Guangdong | 25 |
| Dai 2004(22) | SARS | cross-sectional study | 203.01.18-2003.03.08 | Guangdong | 230 |
| Zhou 2004(23) | SARS | cross-sectional study | to 2003.05 | Guangdong | 2635 |
| Wang 2003(24) | SARS | cross-sectional study | 2003.01.02-2003.04.17 | Guangdong | 966 |
| Gao 2003(25) | SARS | cross-sectional study | 2003.05.14-2003.05.17 | Guangdong | 86 |
| Lin 2003(26) | SARS | cross-sectional study | to 2003.05 | Guangdong | 395 |
| Xu 2003(27) | SARS | cross-sectional study | 2003.01.13-2003.05.05 | Guangdong | 1074 |
| Gao 2003(28) | SARS | cross-sectional study | to 2003.07.07 | / | 669 |
| Yuan 2003(29) | SARS | cross-sectional study | 2003.01-2003.06.20 | Shenzhen | 53 |
| Wang 2003(30) | SARS | cross-sectional study | 2003.04.13-2003.05.08 | Tianjin | 175 |
| Wang 2003(31) | SARS | cross-sectional study | 2003.04.20-2003.05.18 | Tianjin | 2300 |
| Wu 2004(32) | SARS | cross-sectional study | 2003.03.27-2003.06.24 | Beijing | 1861 |
| Huang 2003(33) | SARS | cross-sectional study | 2003.02.02-2002.05 | Guangdong | 454 |
| Li 2003(34) | SARS | cross-sectional study | 2002.12.26-2003.01.19 | Zhongshan | 29 |
| Fei 2003(35) | SARS | cross-sectional study | 2003.03-2003.04 | Beijing | 33 |
| Lu 2003(36) | SARS | case series | from 2003.04.05 | Beijing | 80 |
| He 2003(37) | SARS | cross-sectional study | to 2003.05.20 | Beijing | 2444 |
| Ho 2003(38) | SARS | cross-sectional study | 2003.03.25-05.05 | Hong Kong | 1312 |
| Li 2003(39) | SARS | cross-sectional study | 2003.03.15-05.18 | Beijing | 740 |
| Fowler 2003(40) | SARS | case series | to 2003.04.15 | Tronoto | 38<br>164 |
| Varia 2003(41) | SARS | cross-sectional study | / | Toronto | 128 |
| Lau 2004(42) | SARS | cross-sectional study | / | Hong Kong | 339 |
| Zou 2003(43) | SARS | cross-sectional study | 2003.01.05-05.09 | Guangdong | 1645 |
| Chen 2006(44) | SARS | cross-sectional study | to 2003.07 | Singapore | 105 |
| Cooper 2009(45) | SARS | cross-sectional study | 2003.02.21-03.28 | Beijing | 41 |
|  |  | cross-sectional study | 2003.03.25-04.12 | Beijing | 99 |
|  |  | cross-sectional study | 2003.04.16-05.12 | Tianjin | 91 |
| Oboho 2015(46) | MERS | cross-sectional study | 2014 1.1-5.1 | Saudi Arabia | 255 |
| Xiang 2015(47) | MERS | cross-sectional study | 2015 5.20-7.13 | South Korea | 186 |
| Assiriv 2014(58) | MERS | case series | 2013 4.1-7.12 | Saudi Arabia | 447 |
| Alenazi 2017(49) | MERS | cross-sectional study | 2015 7.15-9.15 | Saudi Arabia | 130 |
| Memish 2015(50) | MERS | cross-sectional study | 2013 8.24-9.3 | Saudi Arabia | 306 |
| Park 2016 (51) | MERS | cross-sectional study | 2015 5.20-7.19 | South Korea | 76 |
|  |  |  |  |  | 70 |
| Al-Dorzi 2016(52) | MERS | case series | 2015 8.25-9.23 | Saudi Arabia | 276 |
| Hunter 2016(53) | MERS | cross-sectional study | 2013 1.1-2014 5.9 | Saudi Arabia | 65 |
| Amer 2018(54) | MERS | cross-sectional study | 2017 3.31 -7 15 | Saudi Arabia | 120 |
| Cho 2016(55) | MERS | case series | 2015 5.27-5.29 | South Korea | 1576 |
| Hijawi 2013 (56) | MERS | cross-sectional study | 2012.4.1-9.30 | Jordan | 13 |

### Nosocomial infections among confirm cases

The proportion of nosocomial infections was 44.0% (95% CI: 0.36 to 0.51; *I*^*2*^=0.00%) among COVID-19 patients, 36.0% (95% CI: 0.23 to 0.49; *I*^*2*^=97.8%) among SARS patients, and 56.0% (95% CI: 0.08 to 1.00; *I*^*2*^=99.9%) among MERS patients (*Figure 2*). Thirty-three percent(95% CI: 0.27 to 0.40; *I*^*2*^=0.00%) of patients with COVID-19 were medical staff, and 2.0% (95% CI: 0.01 to 0.03; *I*^*2*^=0.00%), were nosocomial infections among people other than medical staff (such as inpatients or visitors). The corresponding proportions among SARS patients were 37.0% (95% CI: 0.25 to 0.49; *I*^*2*^=97.3%) and 24.0% (95% CI: 0.10 to 0.38; *I*^*2*^=86.6%), and 19.0% (95% CI: 0.04 to 0.35; *I*^*2*^=97.8%) and 36.0% (95% CI: 0.06 to 0.67; *I*^*2*^=99.3%) among MERS patients (*Figures 3-4*).

**Figure 2.**
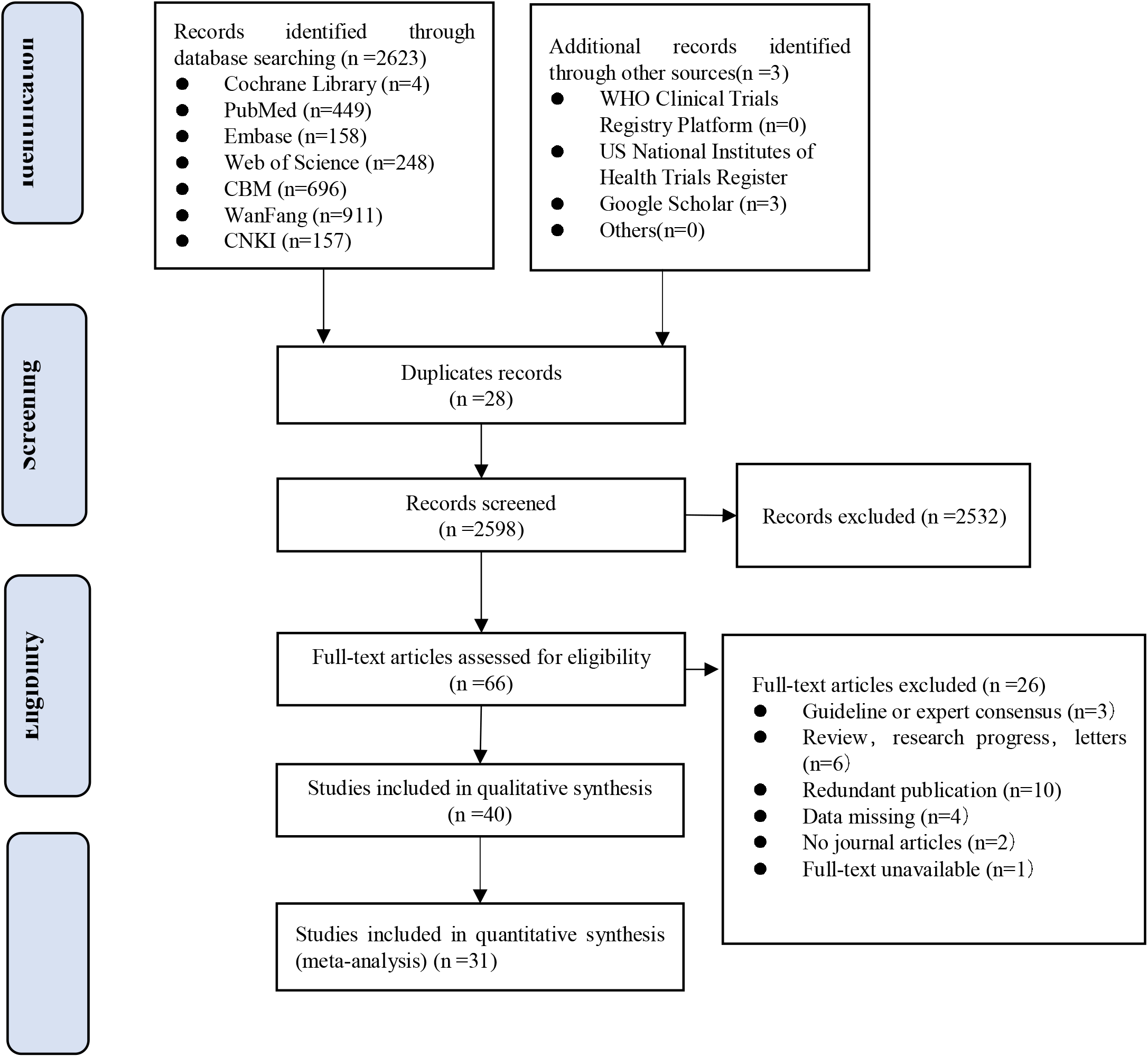
Flow diagram of the literature search.

**Figure 3.**
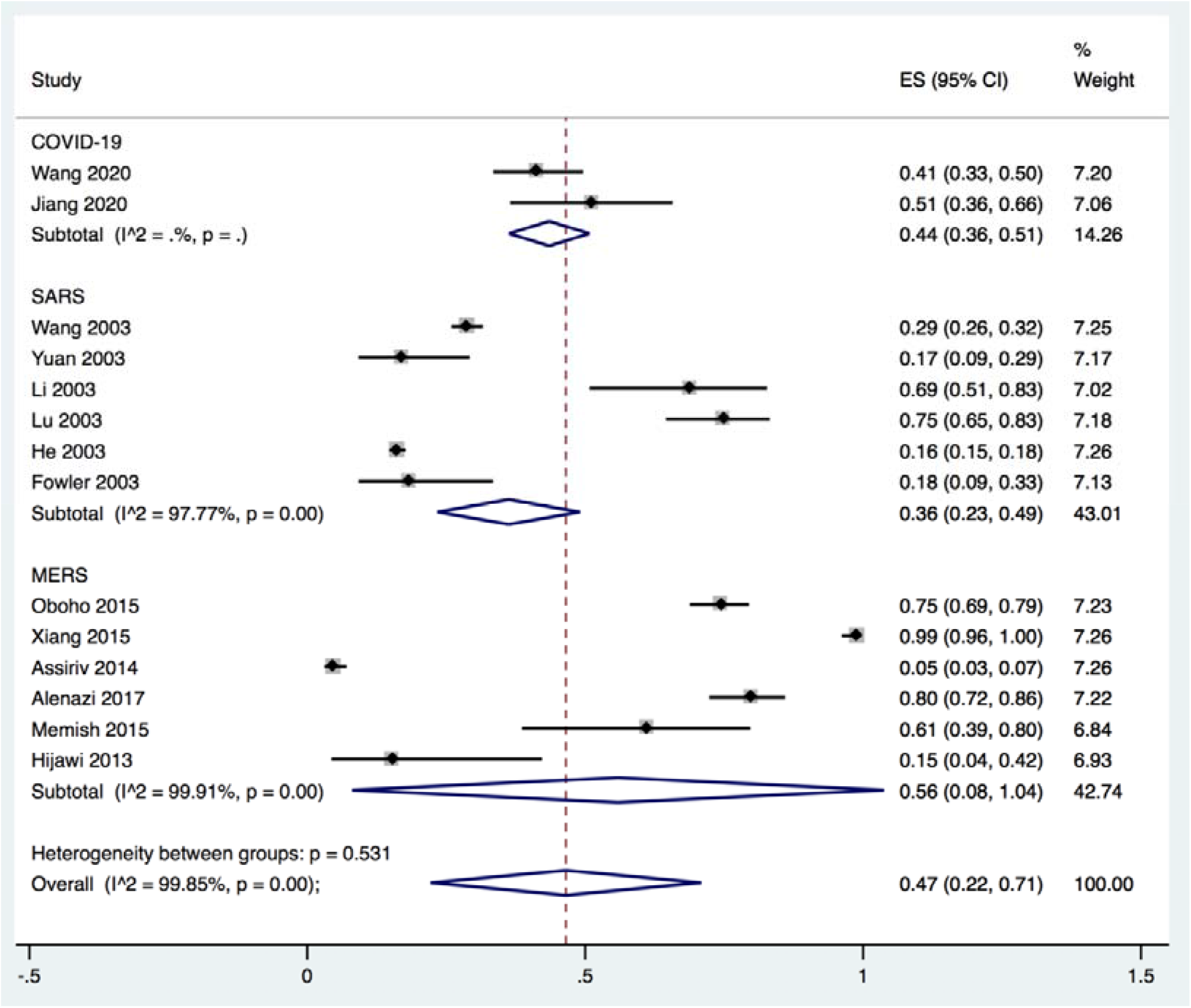
The proportion of nosocomial infections among confirm cases of COVID-19, SARS and MERS.

### Infection among the health care workers

Twenty studies metioned infection among the health workers, of which sixteen studies described the occupational composition of infected health care workers. Doctors accounted for 33.0% (95% CI :0.23 to 0.44), nurses 56.0% (95% CI: 0.45 to 0.66), and other staff (such as carers, cleaners, hospital support staff) 11.0%,(95% CI: 0.06 to 0.20) of COVID-19 cases among hospital staff. For SARS, 30.0% (95% CI:0.19 to 0.40; *I*^*2*^=91.1%) of the cases among hospital workers were doctors, 50.0% (95% CI: 0.45 to 0.55; *I*^*2*^=38.8%) nurses, and 21.0% (95% CI: 0.12 to 0.29; *I*^*2*^=85.2%) others. For MERS, for the corresponding proportions were 35.0% (95% CI:0.14 to 0.56; *I*^*2*^=0.00%), 50.0% (95% CI: 0.29 to 0.71; *I*^*2*^=0.00%) and 16.0% (95% CI: 0.00 to 0.32; *I*^*2*^=0.00%). For all three conditions combined, the proportion of doctors among infected hospital staff was 30.0%, 51.0% for the proportion of nurses, and 19.0% for the proportion of others (*Figure 5-7*).

**Figure 4.**
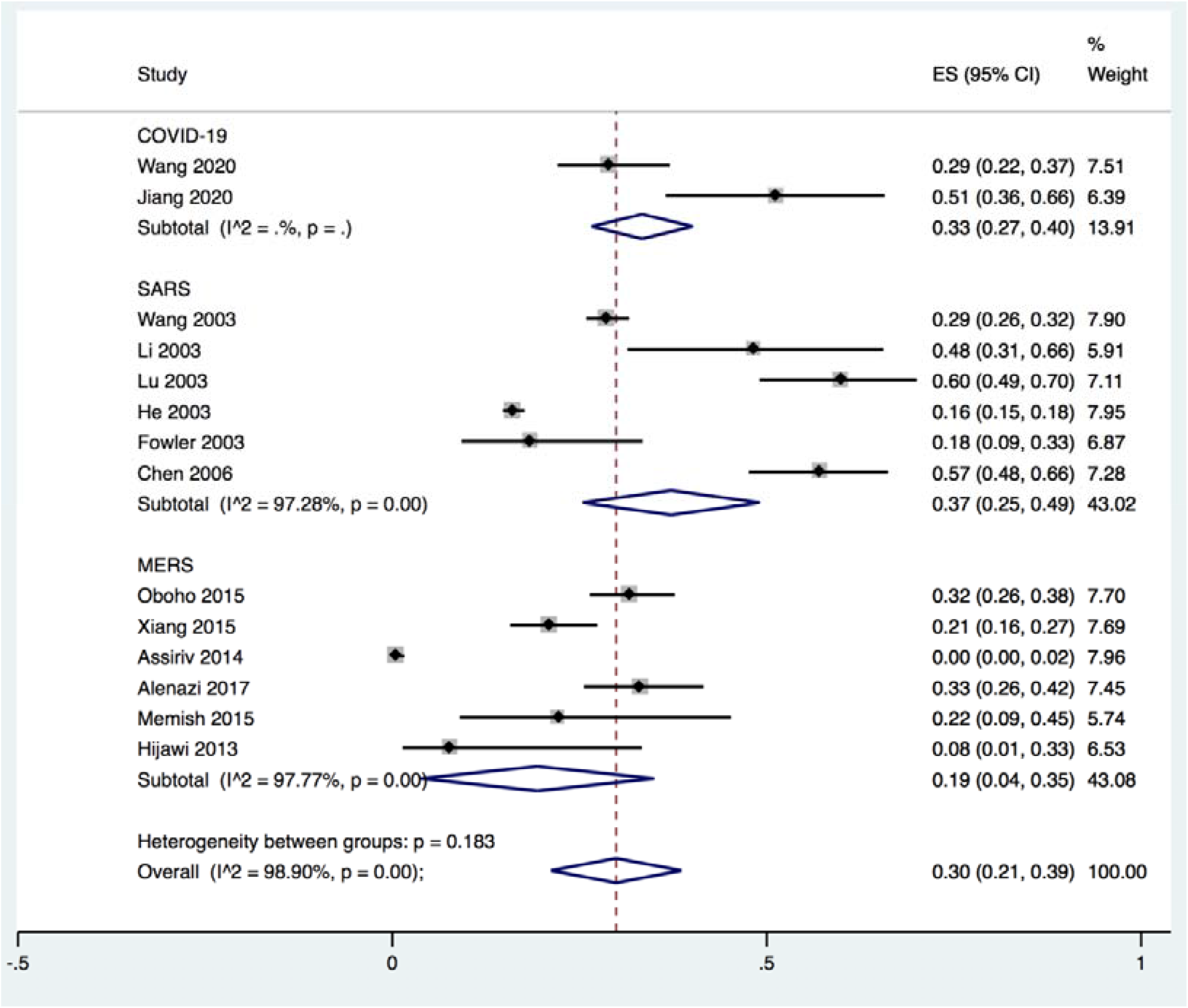
Proportions of health care workers among confirmed cases of COVID-19, SARS and MERS.

**Figure 5.**
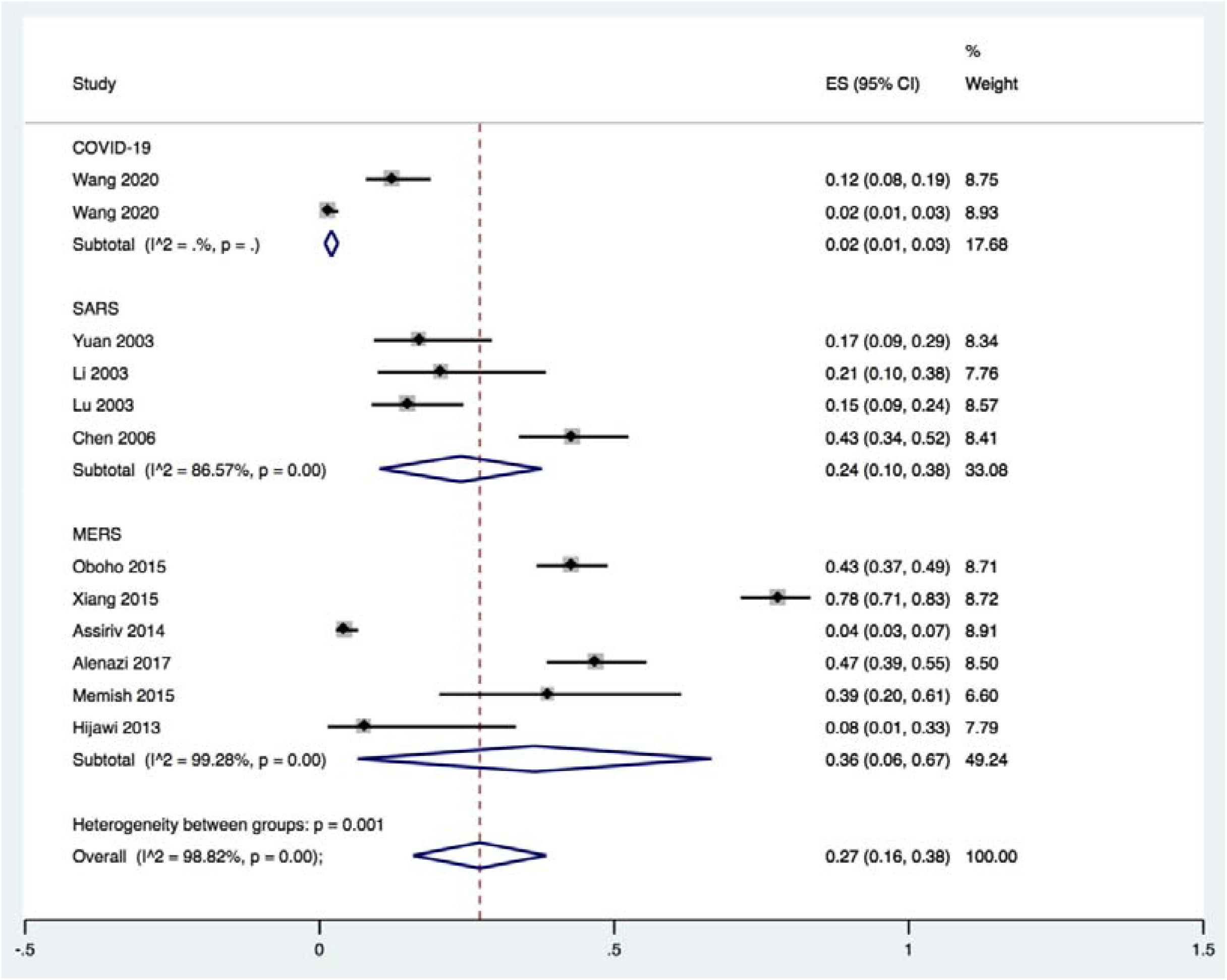
Proportions of nosocomial infections excluding health care workers among confirm cases of COVID-19, SARS and MERS.

Five studies described the protective measures of medical staff infected with SARS in hospital. Sixty-three percent (95% CI: 0.35 to 0.92; *I*^*2*^=96.1%) of the infected staff did not wear protective clothing), 58.0% (95% CI: 0.39 to 0.76; *I*^*2*^=0.00%) did not use gloves, 91.0% (95% CI: 0.80 to 1.00; *I*^*2*^=0.00%) did not wear goggles. 57.0% (95% CI: 0.00 to 1.00; *I*^*2*^=0.00%) did not take any hand disinfection measures), and 7.0% (95% CI: 0.12 to 0.51; *I*^*2*^=0.00%) did not wear masks (*Figure 8*). One study described that among the 22 infected medical workers, 21 had no shoe cover. One study described that of 53 infected health workers, 47 wore cloth masks.

**Figure 6.**
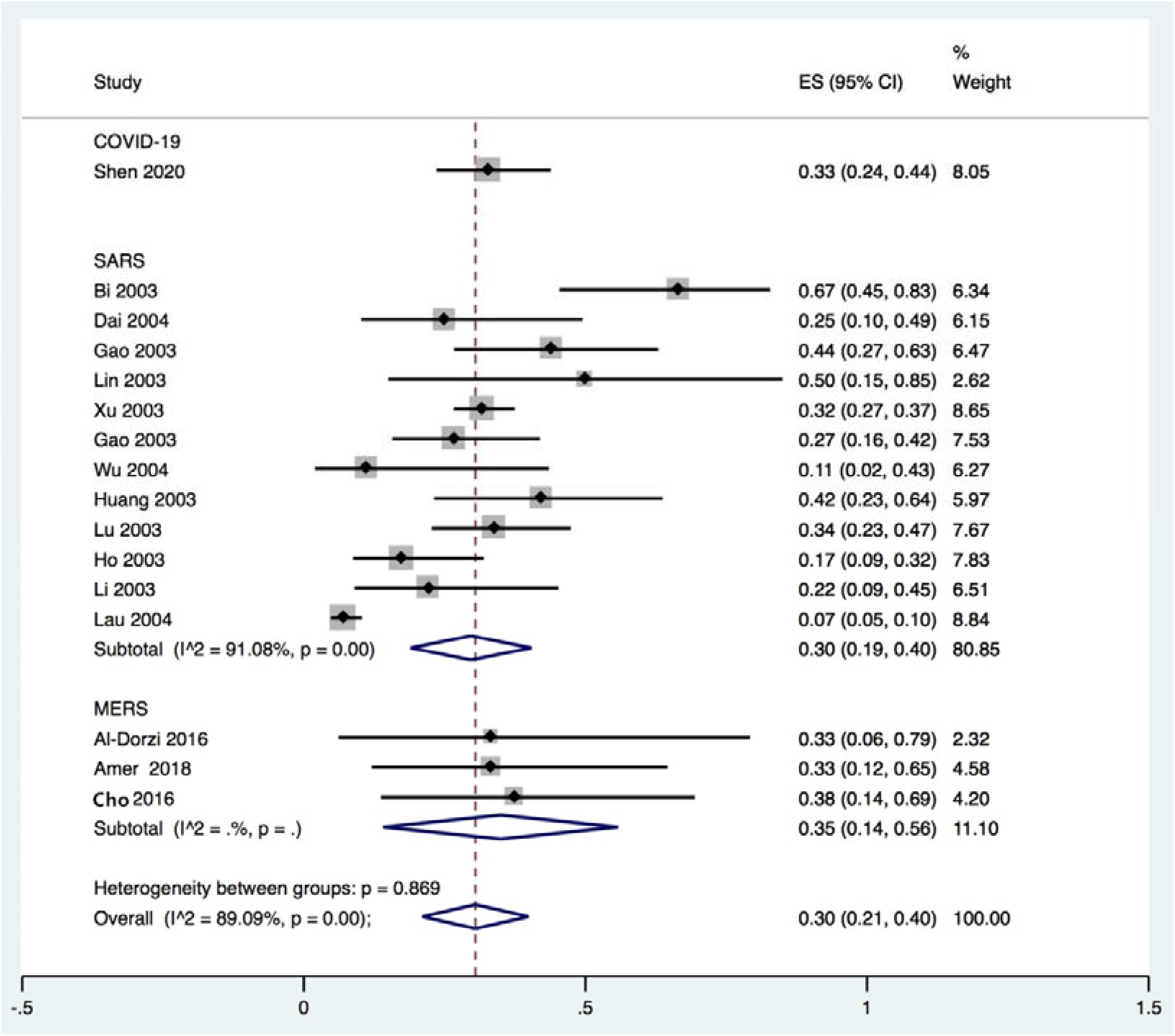
Proportion of doctors among hospital staff with COVID-19, SARS and MERS.

**Figure 7.**
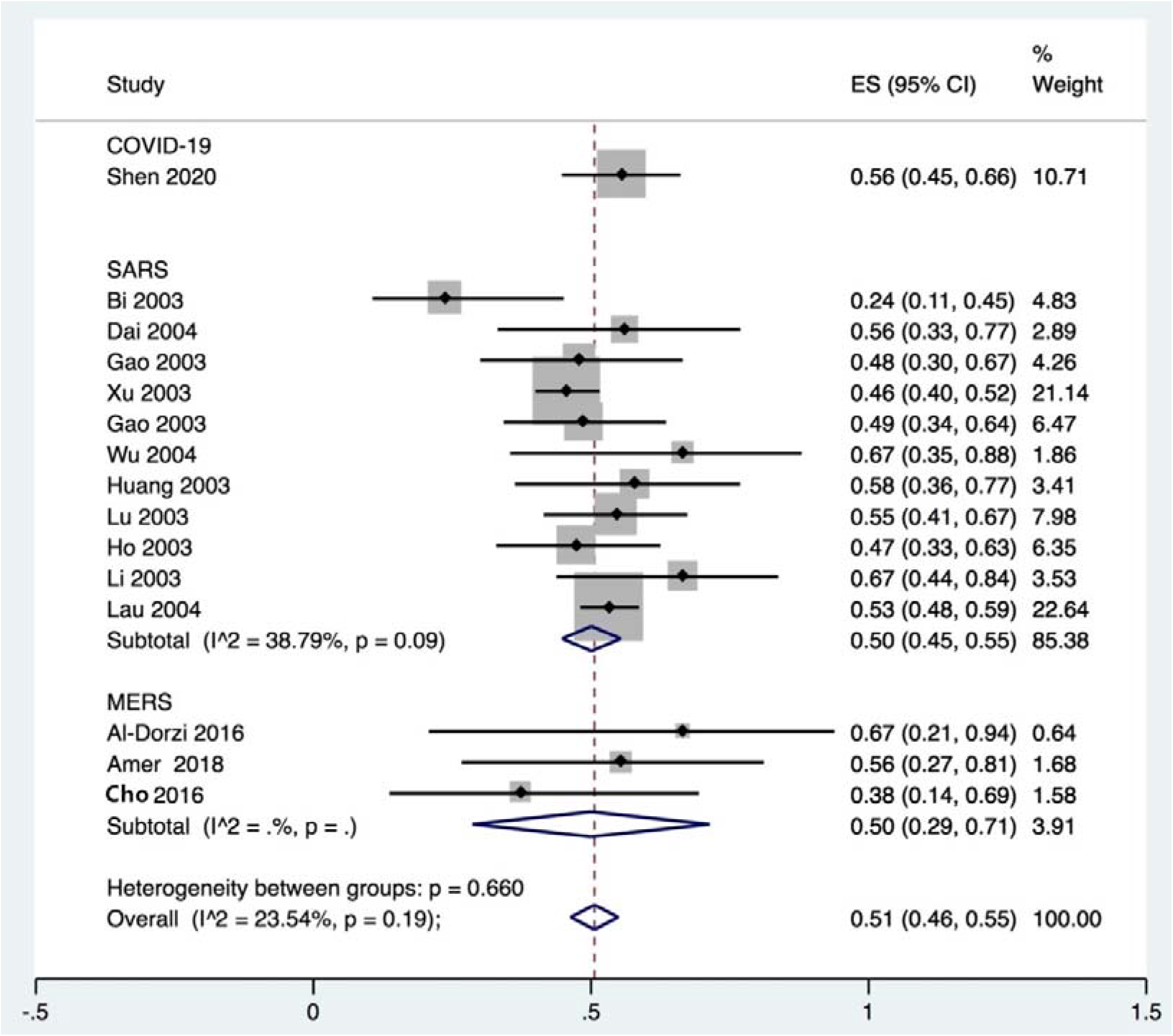
Proportion of nurses among hospital staff with COVID-19, SARS and MERS.

**Figure 8.**
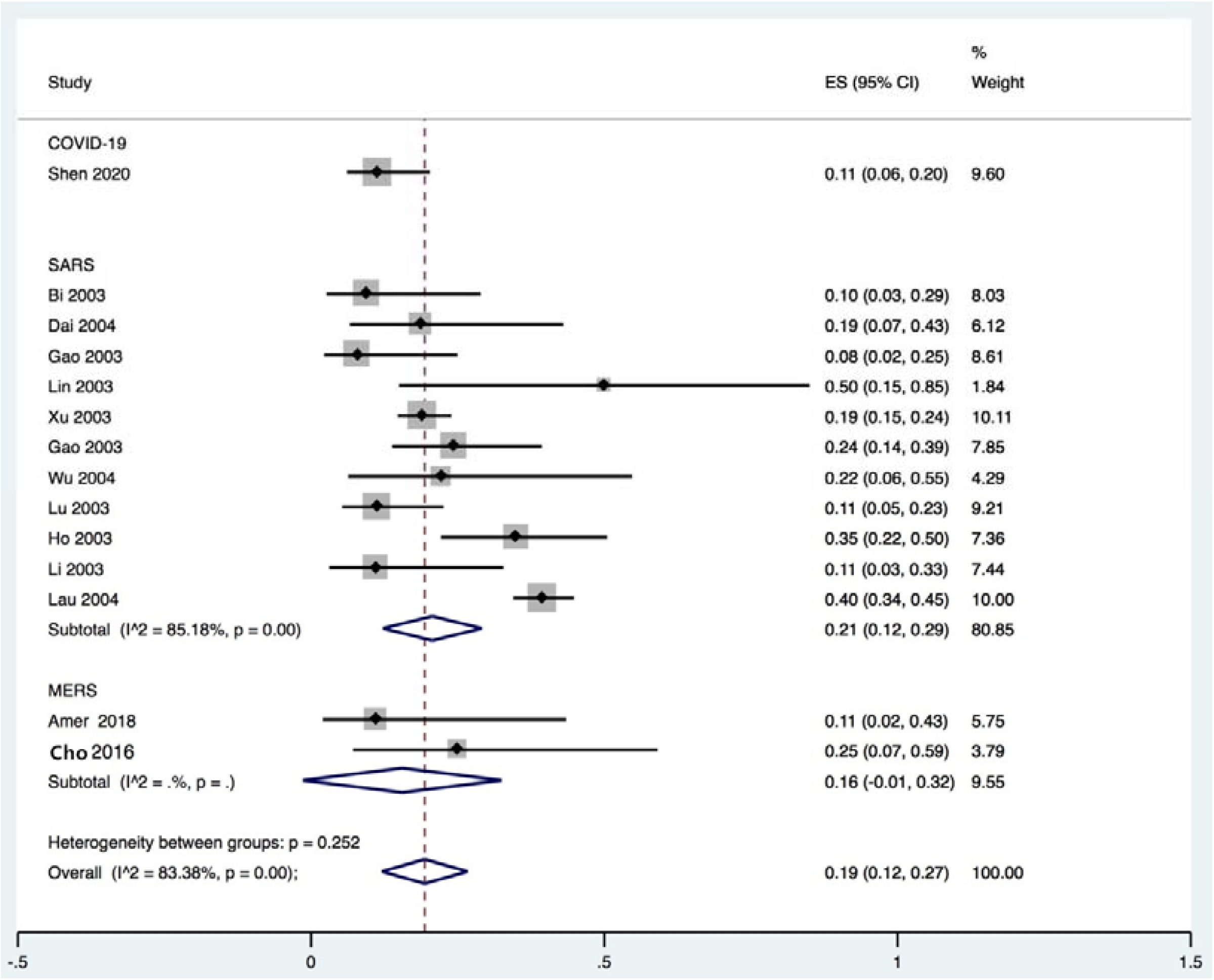
Proportion of staff other than doctors or nurses among hospital staff with COVID-19, SARS and MERS.

**Figure 9.**
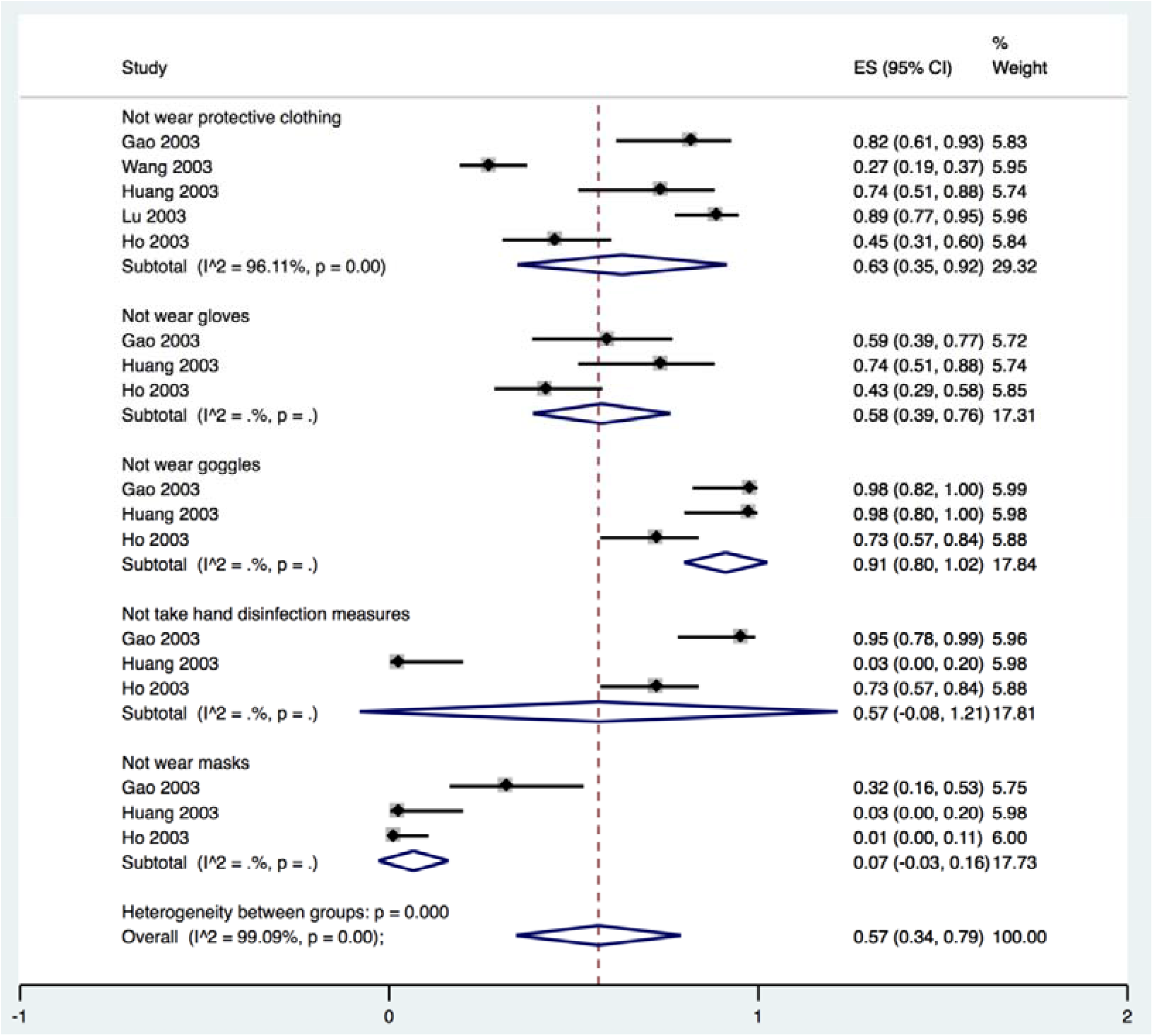
Proportion of health care staff with SARS who did not take protective measures.

### Outbreaks in the hospitals

Six studies described SARS outbreaks, and five studies MERS outbreaks that happened in hospitals. The SARS studies reported on 23 patients, causing a total of 674 infections in hospitals, with an average of 29.3 infections per index patient. The MERS studies reported 24 patients causing 152 infections in hospitals, with an average of 6.3 infections per index patient (*Table 2*).

**Table 2.** Secondary infected by index paitient in outbreaks in the hospitals

| **Study ID** | **Disease** | **Index patients** | **Number of secondary cases** |
| --- | --- | --- | --- |
| Bi 2003(21) | SARS | 3 | 22 |
| Wang 2003(30) | SARS | 1 | 164 |
| Fei 2003(35) | SARS | 2 | 30 |
| Varia 2003(41) | SARS | 6 | 126 |
| Chen 2006(44) | SARS | 7 | 105 |
| Cooper 2009(45) | SARS | 4 | 227 |
| **Total** |  | 23 | 674 |
| Memish 2015(50) | MERS | 18 | 4 |
| Park 2016(51) | MERS | 1 | 23 |
| Hunter 2016(53) | MERS | 3 | 27 |
| Amer 2018(54) | MERS | 1 | 16 |
| Cho 2016(55) | MERS | 1 | 82 |
| **Total** |  | 24 | 152 |

### Quality of evidence

The results of GRADE on nosocomial infections showed that the quality of evidence were low or very low. The details can be found in the **Supplementary Material 3**.

## Discussion

Our rapid review identified a total of 40 studies. Low to very low-quality evidence indicated that the proportion of nosocomial infection among confirmed cases of COVID-19 was 44%, which is higher than for SARS but lower than for MERS. Most patients with COVID-19 and SARS infected in hospitals were medical staff, among whom nurses formed the largest group, followed by doctors. Both SARS and MERS outbreaks have been reported in hospitals, but we found no evidence of a COVID-19 outbreak.

SARS-CoV-2, the infectious agent causing COVID-19, is highly contagious, mainly spread by droplets and close contact. So far, a number of familial disease clusters have been reported, and some of the confirmed patients had been infected in healthcare facilities. As health care workers are in contact with a large number of suspected patients on a daily basis, strict precautions need to be taken to avoid outbreaks of infection in health care facilities. In the early stage of the epidemic, some hospitals and staff did not have enough knowledge about the virus, leading to inadequate prevention and control measures. Suspected patients did often not take any protection measures when they went to the hospital, which may have caused nosocomial infections and hospital outbreaks(19,20). A MERS study showed routine infection-prevention policies can greatly reduce nosocomial transmission of MERS(57). According to a report by the WHO, 20% of confirmed cases of SARS were among health care workers(58). Due to the rapidly evolving outbreak and spread of the disease, medical staff need to work in a state of high tension, but they should also protect themselves adequately and take the appropriate isolation measures to avoid cross infection in the hospital. The high presence of the COVID-19 epidemic in the media is likely to improve the general public’s awareness. People with symptoms indicating a SARS-CoV-2 infection should take protective measures during the hospital or clinic visit, such as wearing a mask, minimizing the time of stay in the hospital, and if possible, making remote medical consultation in advance. Medical institutions should formulate sound infection prevention and control strategies, and strengthen the hospital’s infection prevention and control efforts, such as the establishment of special departments for outpatients with fever, and a sound triage system: triage of early identification among suspected cases can avoid excessive gathering of patients in the hospital. Isolation wards should be established for suspected and confirmed patients needing treatment. In hospitals without single isolation wards or negative pressure isolation, indoor ventilation measures should be taken timely, and the management of patients should be standardized in these wards. Using adequate disinfection procedures can reduce the possibility of hospital transmission of the virus. During the epidemic, efforts should be made to publicize the knowledge of infection prevention and control, be alert to the possibility of the outbreak of nosocomial infection, and establish an early warning mechanism. Emergency plans or measures should be developed to deal with nosocomial infections.

### Strengths and Limitations

Our study included studies related to nosocomial infections among COVID-19, SARS and MERS patients. Our results can help the decision-making related to prevention, control and clinical management in hospitals. Some studies had missing data, and we used methods of meta-analyses of proportions to analyse those studies with available data, so the proportions estimated may not be accurate and similar to the actual data. Most of the results are based on low-quality research, so that the credibility of the results is low.

## Conclusion

A large proportion of confirmed cases of COVID-19 were infected within healthcare facilities. Therefore, the patients who come to the hospital should do pay attention on personal protection. At the same time, medical institutions can reduce the spread of the virus through triage, and setting up separate fever clinic and isolation wards. Awareness of the disease needs to be improved among medical staff, so that they can protect themselves adequately and stop the spread of the virus within hospitals.

## Data Availability

This is a rapid review. All data were from the original and published studies.

## Author contributions

(I) Conception and design: Y Chen and E Liu; (II) Administrative support: Y Chen; (III) Provision of study materials or patients: Y Gao, R Liu and X Wang; (IV) Collection and assembly of data: R Liu, X Wang, YL Gao, P DU, X Wang, X Zhang, S Lu and Z Wang; (V) Data analysis and interpretation: Q Zhou, Q Shi and Y Gao; (VI) Manuscript writing: All authors; (VII) Final approval of manuscript: All authors.

## Acknowledgments

We thank Janne Estill, Institute of Global Health of University of Geneva for providing guidance and comments for our review. We thank all the authors for their wonderful collaboration.

## Footnote

### Ethical Statement

The authors are accountable for all aspects of the work in ensuring that questions related to the accuracy or integrity of any part of the work are appropriately investigated and resolved.

## Supplementary Material 1-Search strategy

### PubMed

#1 “COVID-19”[Supplementary Concept]

#2 “Severe Acute Respiratory Syndrome Coronavirus 2”[Supplementary Concept]

#3 “Middle East Respiratory Syndrome Coronavirus”[Mesh]

#4 “Severe Acute Respiratory Syndrome”[Mesh]

#5 “SARS Virus”[Mesh]

#6 “COVID-19”[Title/Abstract]

#7 “SARS-COV-2”[Title/Abstract]

#8 “Novel coronavirus”[Title/Abstract]

#9 “2019-novel coronavirus”[Title/Abstract]

#10 “coronavirus disease-19”[Title/Abstract]

#11 “coronavirus disease 2019”[Title/Abstract]

#12 “COVID19”[Title/Abstract]

#13 “Novel CoV”[Title/Abstract]

#14 “2019-nCoV”[Title/Abstract]

#15 “2019-CoV”[Title/Abstract]

#16 “Wuhan-Cov”[Title/Abstract]

#17 “Wuhan Coronavirus” [Title/Abstract]

#18 “Wuhan seafood market pneumonia virus”[Title/Abstract]

#19 “Middle East Respiratory Syndrome”[Title/Abstract]

#20 “MERS”[Title/Abstract]

#21 “MERS-CoV”[Title/Abstract]

#22 “Severe Acute Respiratory Syndrome”[Title/Abstract]

#23 “SARS”[Title/Abstract]

#24 “SARS-CoV”[Title/Abstract]

#25 “SARS-Related”[Title/Abstract]

#26 “SARS-Associated”[Title/Abstract]

#27#1-#26/ OR

#28 “Cross Infection”[MeSH Terms]

#29 “Cross Infection*”[Title/Abstract]

#30 “Healthcare Associated Infections*”[Title/Abstract]

#31 “Health Care Associated Infection*”[Title/Abstract]

#32 “Hospital Infection*”[Title/Abstract]

#33 “Nosocomial Infection*”[Title/Abstract]

#34 “hospital-related infection*”[Title/Abstract]

#35 “hospital-acquired infection*”[Title/Abstract]

#36#28-# 35/OR

#37#27 AND#36

### Embase

#1 ‘middle east respiratory syndrome coronavirus’/exp

#2 ‘severe acute respiratory syndrome’/exp

#3 ‘sars coronavirus’/exp

#4 ‘COVID-19’:ab,ti

#5 ‘SARS-COV-2’:ab,ti

#6 ‘novel coronavirus’:ab,ti

#7 ‘2019-novel coronavirus’:ab,ti

#8 ‘coronavirus disease-19’:ab,ti

#9 ‘coronavirus disease 2019’:ab,ti

#10 ‘COVID19’:ab,ti

#11 ‘novel cov’:ab,ti

#12 ‘2019-ncov’:ab,ti

#13 ‘2019-cov’:ab,ti

#14 ‘wuhan-cov’:ab,ti

#15 ‘wuhan coronavirus’:ab,ti

#16 ‘wuhan seafood market pneumonia virus’:ab,ti

#17 ‘middle east respiratory syndrome’:ab,ti

#18 ‘middle east respiratory syndrome coronavirus’:ab,ti

#19 ‘mers’:ab,ti

#20 ‘mers-cov’:ab,ti

#21 ‘severe acute respiratory syndrome’:ab,ti

#22 ‘sars’:ab,ti

#23 ‘sars-cov’:ab,ti

#24 ‘sars-related’:ab,ti

#25 ‘sars-associated’:ab,ti

#26#1-#25/ OR

#27 ‘hospital infection*’:ab,ti

#28 ‘nosocomial infection*’:ab,ti

#29 ‘hospital-related infection*’:ab,ti

#30 ‘hospital-acquired infection*’:ab,ti

#31 ‘cross infection*’:ab,ti

#32 ‘healthcare associated infection*’:ab,ti

#33 ‘health care associated infection*’:ab,ti

#34#27-#33/ OR

#35#26 AND#34

### Web of science

#1 TOPIC: “COVID-19”

#2 TOPIC: “SARS-COV-2”

#3 TOPIC: “Novel coronavirus”

#4 TOPIC: “2019-novel coronavirus”

#5 TOPIC: “coronavirus disease-19”

#6 TOPIC: “coronavirus disease 2019”

#7 TOPIC: “COVID 19”

#8 TOPIC: “Novel CoV”

#9 TOPIC: “2019-nCoV”

#10 TOPIC: “2019-CoV”

#11 TOPIC: “Wuhan-Cov”

#12 TOPIC: “Wuhan Coronavirus”

#13 TOPIC: “Wuhan seafood market pneumonia virus”

#14 TOPIC: “Middle East Respiratory Syndrome”

#15 TOPIC: “MERS”

#16 TOPIC: “MERS-CoV”

#17 TOPIC: “Severe Acute Respiratory Syndrome”

#18 TOPIC: “SARS”

#19 TOPIC: “SARS-CoV”

#20 TOPIC: “SARS-Related”

#21 TOPIC: “SARS-Associated”

#22#1-#21/OR

#23 TITLE: “Healthcare Associated Infection”

#24 TITLE: “Healthcare Associated Infections”

#25 TITLE: “Health Care Associated Infection”

#26 TITLE: “Health Care Associated Infections”

#27 TITLE: “Hospital Infection”

#28 TITLE: “Nosocomial Infection”

#29 TITLE: “Nosocomial Infections”

#30 TITLE: “Hospital Infections”

#31 TITLE: “hospital-related infection”

#32 TITLE: “hospital-acquired infection”

#33 TITLE: “Cross Infection”

#34 TITLE: “Cross Infections”

#35#35.#23-#34/OR

#36#36.#22 AND#35

### Cochrane library

#1 MeSH descriptor: [Middle East Respiratory Syndrome Coronavirus] explode all trees

#2 MeSH descriptor: [Severe Acute Respiratory Syndrome] explode all trees

#3 MeSH descriptor: [SARS Virus] explode all trees

#4 “COVID-19”:ti,ab,kw

#5 “SARS-COV-2”:ti,ab,kw

#6 “Novel coronavirus”:ti,ab,kw

#7 “2019-novel coronavirus” :ti,ab,kw

#8 “Novel CoV” :ti,ab,kw

#9 “2019-nCoV” :ti,ab,kw

#10 “2019-CoV” :ti,ab,kw

#11 “coronavirus disease-19” :ti,ab,kw

#12 “coronavirus disease 2019” :ti,ab,kw

#13 “COVID19” :ti,ab,kw

#14 “Wuhan-Cov” :ti,ab,kw

#15 “Wuhan Coronavirus” :ti,ab,kw

#16 “Wuhan seafood market pneumonia virus” :ti,ab,kw

#17 “Middle East Respiratory Syndrome” :ti,ab,kw

#18 “MERS”:ti,ab,kw

#19 “MERS-CoV”:ti,ab,kw

#20 “Severe Acute Respiratory Syndrome”:ti,ab,kw

#21 “SARS” :ti,ab,kw

#22 “SARS-CoV” :ti,ab,kw

#23 “SARS-Related”:ti,ab,kw

#24 “SARS-Associated”:ti,ab,kw

#25#1-#24/ OR

#26 “hospital-related infection*”:ti,ab,kw

#27 “hospital-related infection*”:ti,ab,kw

#28 “cross infection*”:ti,ab,kw

#29 “healthcare associated infection*”:ti,ab,kw

#30 “health care associated infection*”:ti,ab,kw

#31 “hospital infection*”:ti,ab,kw

#32 “nosocomial infection*”:ti,ab,kw

#33#26-#32/OR

#34#34.#25 AND#33

### CNKI

#1 “新型冠状病毒”[主题]

#2 “COVID-19”[主题]

#3 “COVID 19”[主题]

#4 “2019-nCoV”[主题]

#5 “2019-CoV”[主题]

#6 “SARS-CoV-2”[主题]

#7 “武汉冠状病毒”[主题]

#8 “中东呼吸综合征”[主题]

#9 “MERS”[主题]

#10 “MERS-CoV”[主题]

#11 “严重急性呼吸综合征”[主题]

#12 “SARS”[主题]

#13 #1-#12/ OR

#14 “医院相关感染” [主题]

#15 “医院获得性感染” [主题]

#16 “医疗机构相关感染” [主题]

#17 “院内感染” [主题]

#18 “交叉感染” [主题]

#19 #14-#18/ OR

#20 #13 AND#19

### WanFang

#1 “新型冠状病毒”[主题]

#2 “COVID-19”[主题]

#3 “COVID 19”[主题]

#4 “2019-nCoV”[主题]

#5 “2019-CoV”[主题]

#6 “SARS-CoV-2”[主题]

#7 “武汉冠状病毒”[主题]

#8 “中东呼吸综合征”[主题]

#9 “MERS”[主题]

#10 “MERS-CoV”[主题]

#11 “严重急性呼吸综合征”[主题]

#12 “SARS”[主题]

#13 #1-#12/ OR

#14 “医院相关感染”)[主题]

#15 (“医院获得性感染”)[主题]

#16 (“医疗机构相关感染”)[主题]

#17 (“院内感染”)[主题]

#18 (“交叉&#x感染”)[主题]

#19 #14-#18/OR

#20 #18 AND #19

### CBM

#1 “新型冠状病毒”[常用字段:智能]

#2 “COVID-19”[常用字段:智能]

#3 “COVID 19”[常用字段:智能]

#4 “2019-nCoV”[常用字段:智能]

#5 “2019-CoV”[常用字段:智能]

#6 “SARS-CoV-2”[常用5B57段:智能]

#7 “武汉冠状病毒”[常用字段:智能]

#8 “中东呼吸综合征冠状病毒”[不加权:扩展]

#9 “中东呼吸综合征”[常用字段:智能]

#10 “MERS”[常用字段:智能]

#11 “MERS-CoV”[常用字段:智能]

#12 “严重急性呼吸综合征”[不加权:扩展]

#13 “SARS 病毒”[不加权:扩展]

#14 “严重急性呼吸综合征”[常用字段:智能]

#15 “SARS”[常用字段:智能]

#16 #1-#15/OR

#17 “医院相关感染”[常用字段:智能]

#18 “医院获得性感染”[常用字段:智能]

#19 “医疗机构相关感染”[常;用字段:智能]

#20 “交叉感染”[常用字段:智能]

#21 “院内感染”[常用字段:智能]

#22 #17-#21/OR

#23#16 AND#22

## Supplementary Material 2- Risk of bias in the included studies

**Table A:**
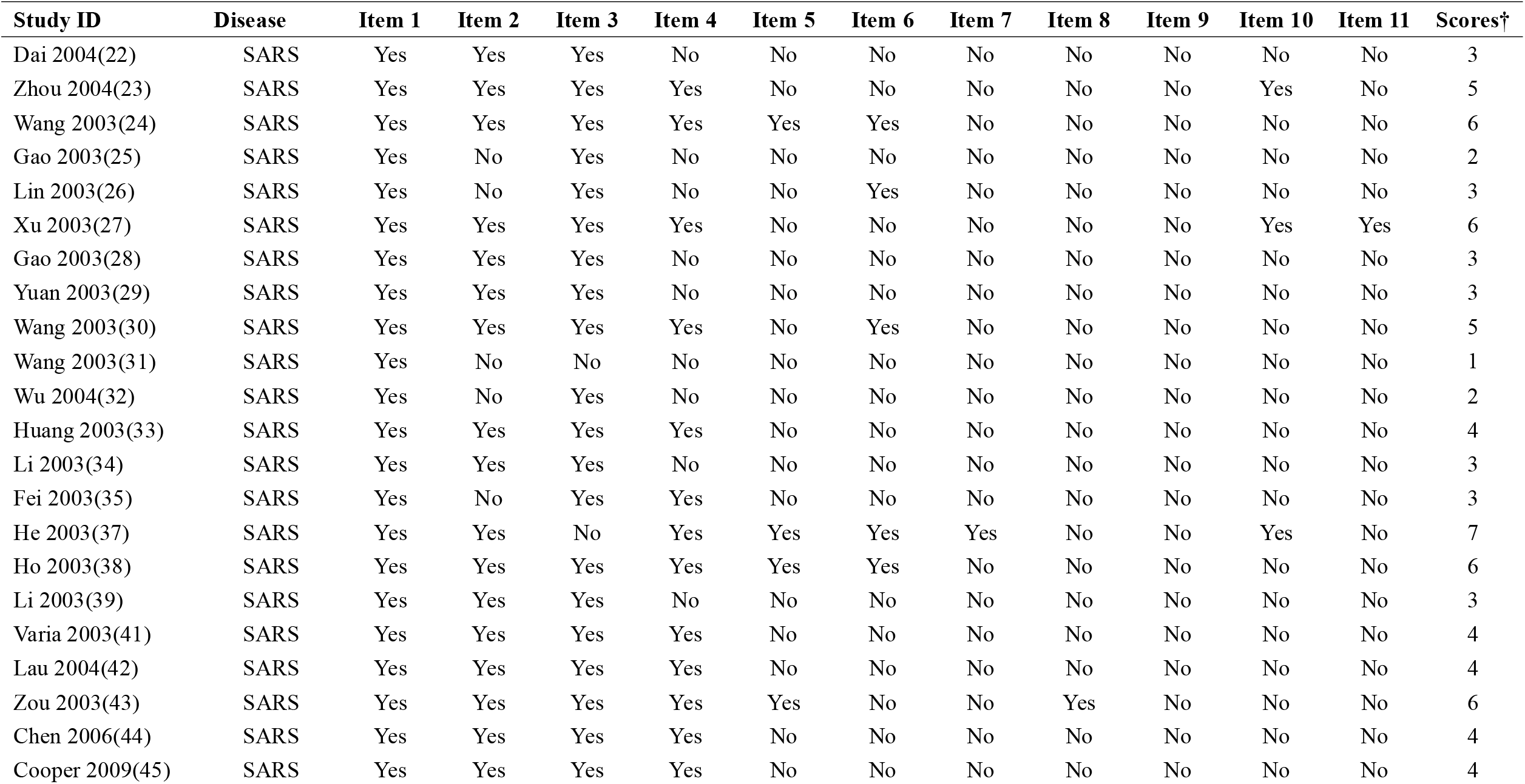
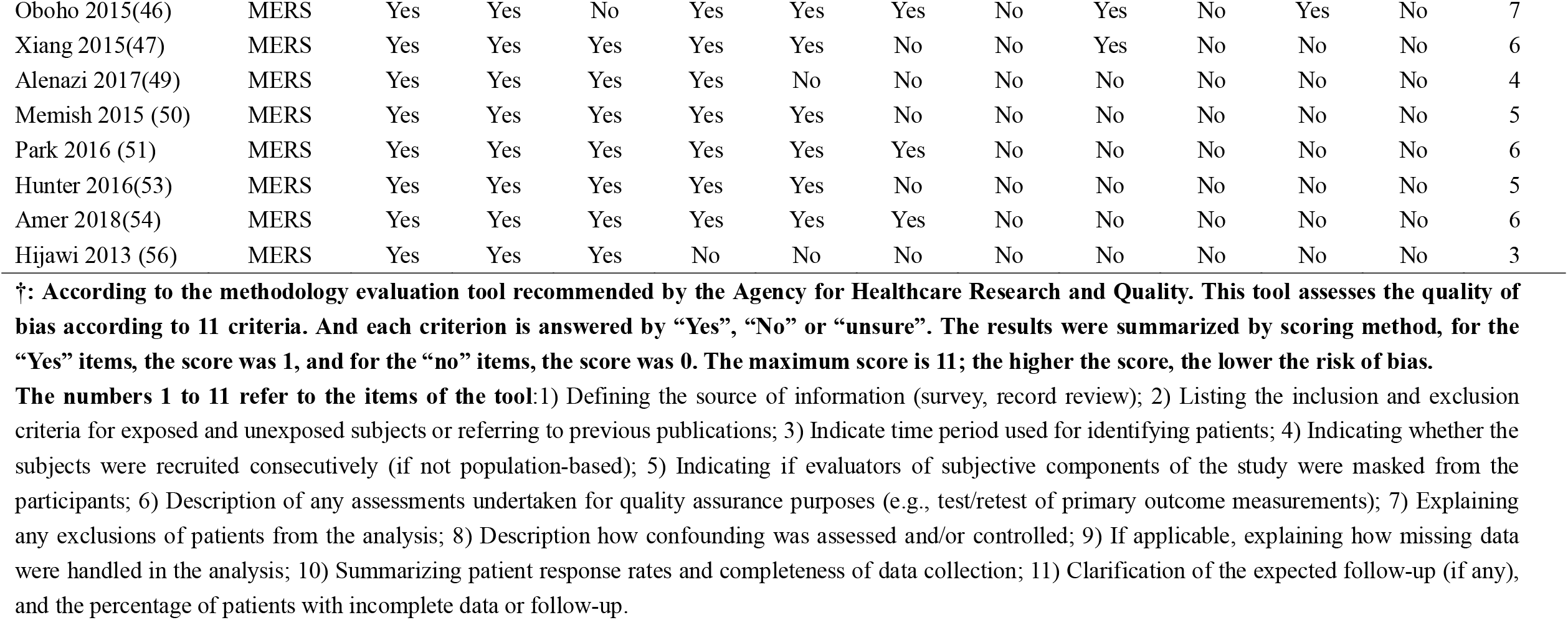
Cross-sectional studies

**Table B:** Case series

| Study ID | Disease | Item 1 | Item 2 | Item 3 | Item 4 | Item 5 | Item 6 | Item 7 | Item 8 | Scores†† |
| --- | --- | --- | --- | --- | --- | --- | --- | --- | --- | --- |
| Wang 2020(8) | COVID-19 | Yes | Yes | Yes | No | No | Yes | Yes | Yes | 6 |
| Wang 2020(18) | COVID-19 | No | Yes | No | No | No | Yes | Yes | Yes | 4 |
| Jiang 2020(19) | COVID-19 | Yes | Yes | Yes | No | No | Yes | Yes | Yes | 6 |
| Bi 2003(21) | SARS | No | Yes | No | No | No | No | Yes | Yes | 3 |
| Lu 2003(36) | SARS | No | Yes | No | No | No | No | Yes | Yes | 3 |
| Fowler 2003(40) | SARS | Yes | Yes | Yes | Yes | No | No | Yes | Yes | 6 |
| Assiriv 2014(58) | MERS | Yes | Yes | Yes | No | No | No | Yes | Yes | 5 |
| Al-Dorzi 2016(52) | MERS | No | Yes | Yes | No | No | No | Yes | Yes | 4 |
| Cho 2016(55) | MERS | Yes | Yes | Yes | Yes | No | No | Yes | Yes | 6 |

††: According to the methodology evaluation tool recommended by National Institute for Health and Care Excellence. The risk of bias is evaluated according to eight criteria. The results were summarized by scoring method, for the “Yes” items, the score was 1, and for the “no” items, the score was 0. The maximum score is 8; the higher the score, the lower the risk of bias.
The numbers 1 to 8 refer to the items of the tool: 1) Case series collected in more than one centre, i.e. multi-centre study; 2) Is the hypothesis/aim/objective of the study clearly described?; 3) Are the inclusion and exclusion criteria (case definition) clearly reported?; 4) Is there a clear definition of the outcomes reported?; 5) Were data collected prospectively?; 6) Is there an explicit statement that patients were recruited consecutively?; 7) Are the main findings of the study clearly described?; 8) Are outcomes stratified? (e.g., by disease stage, abnormal test results, patient characteristics)

**Table C:**
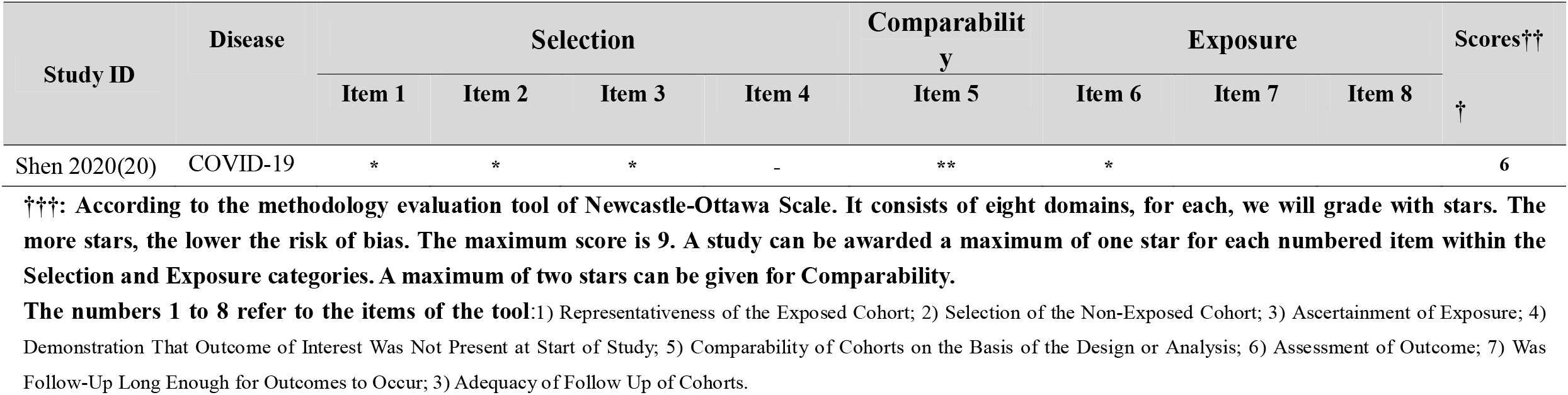
Case control study

## Supplementary Material 3-Summary of Findings

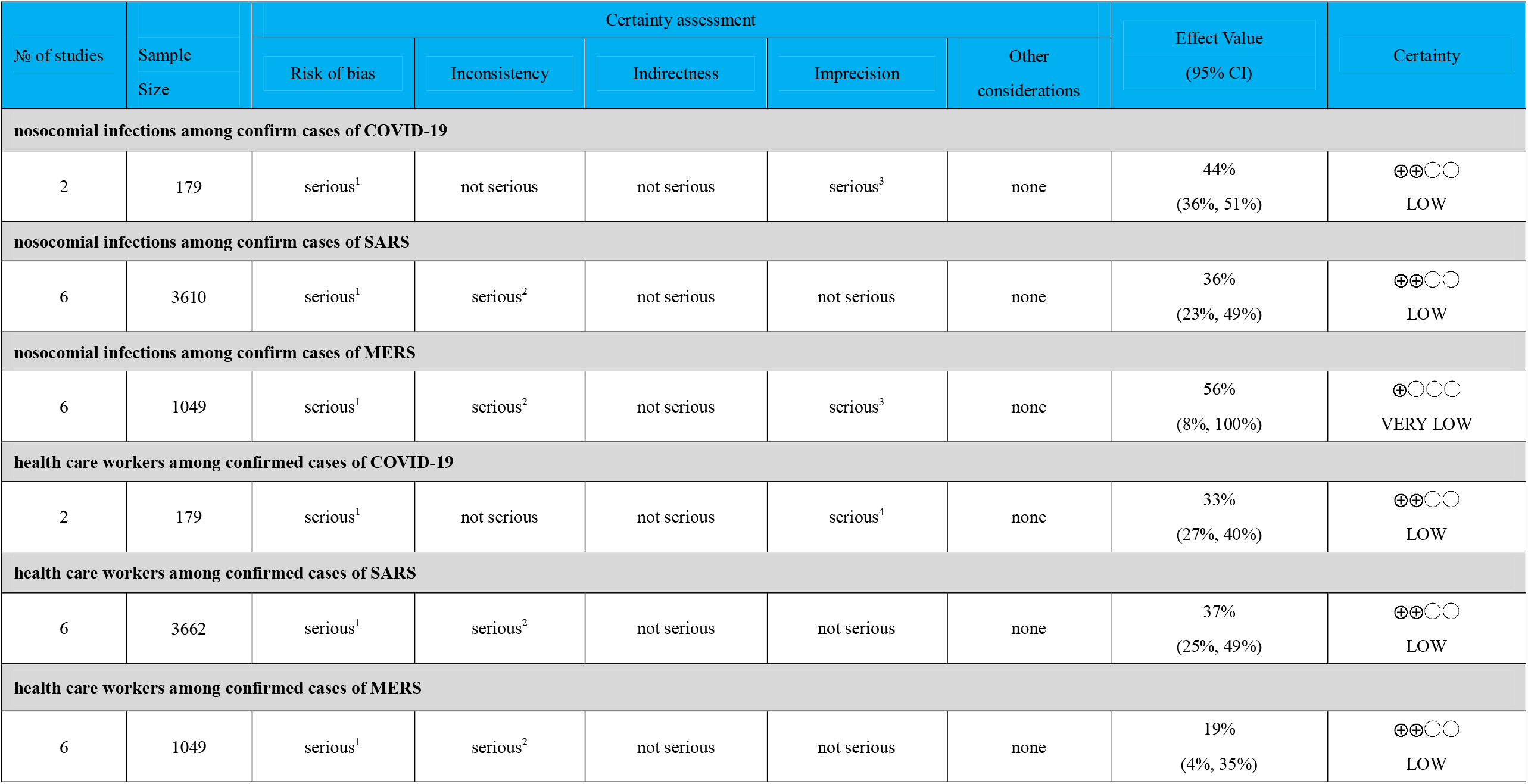
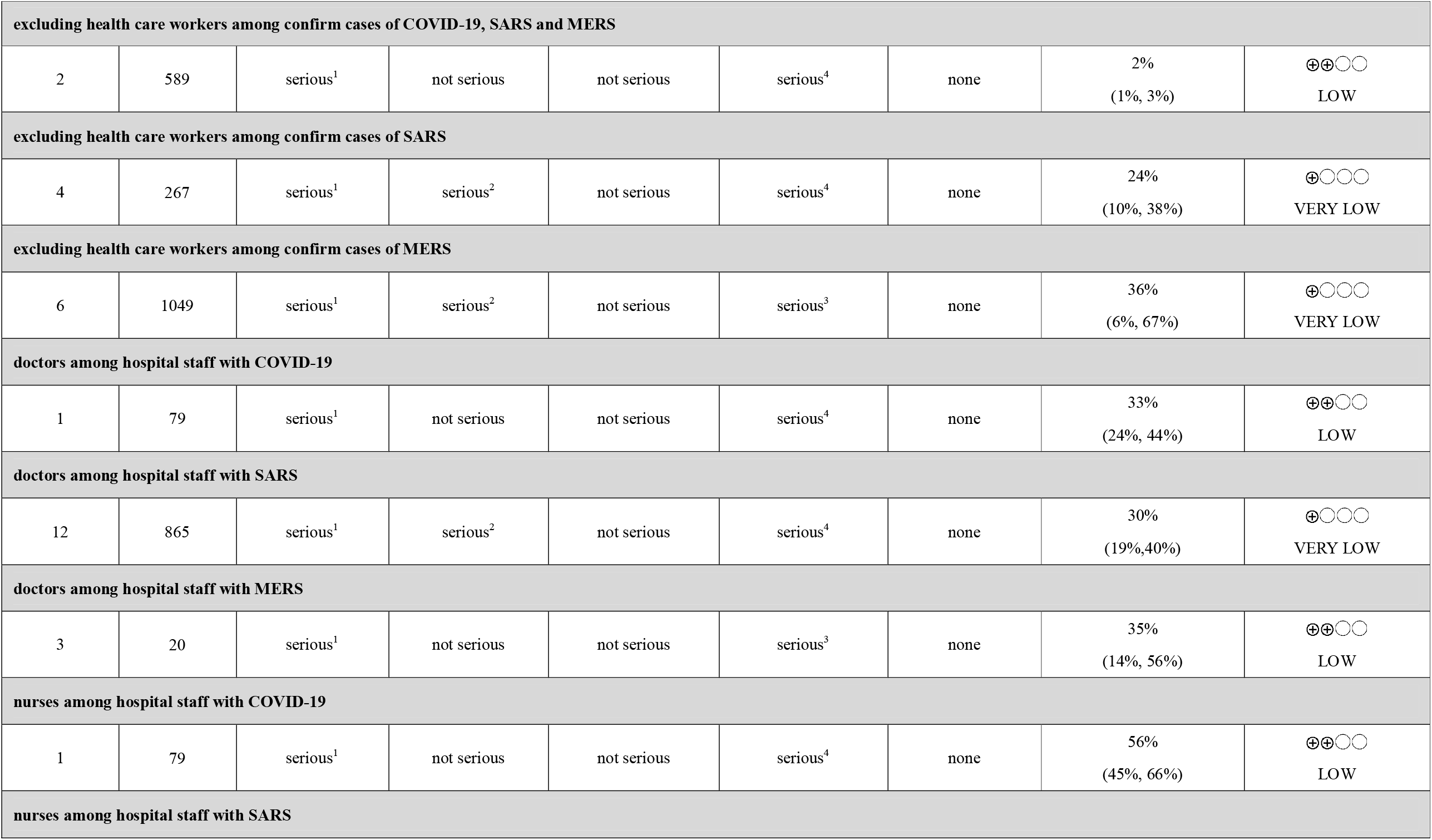
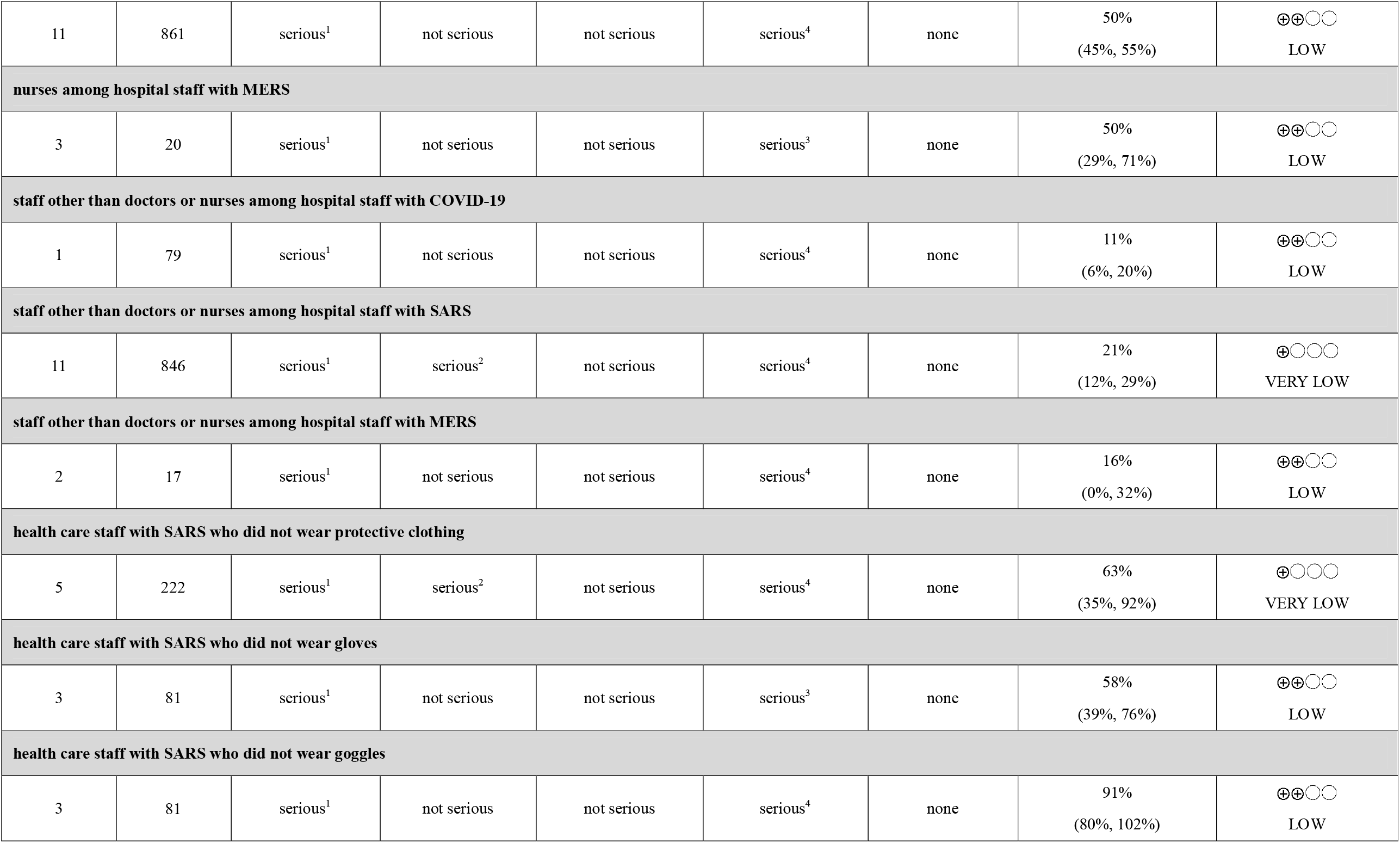
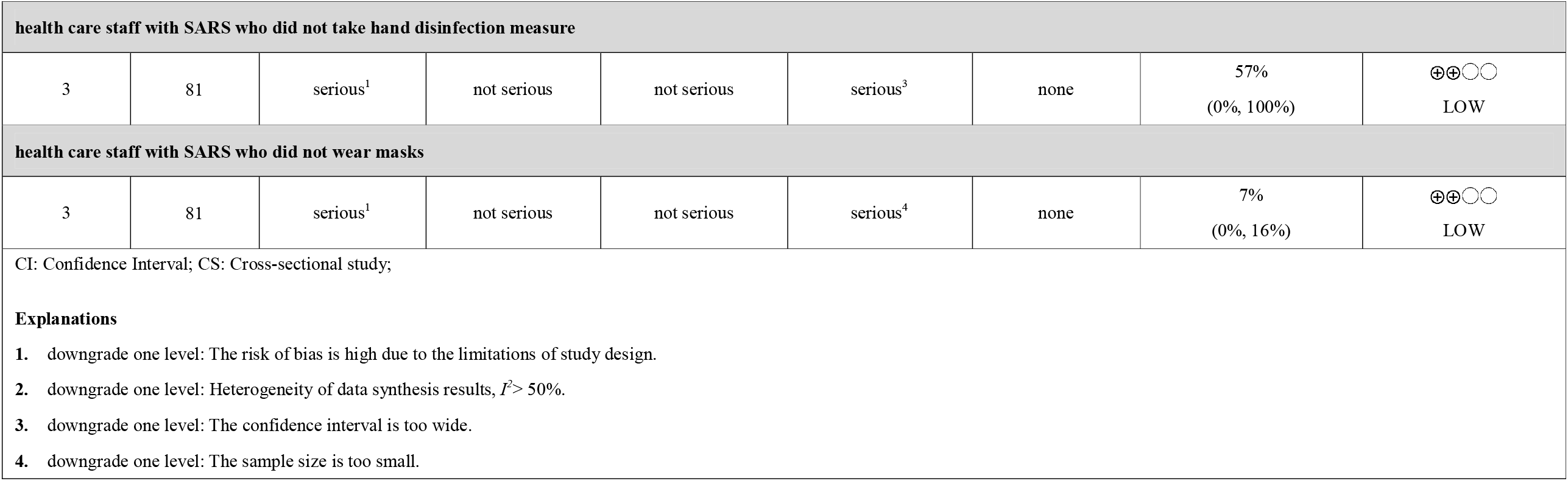

## Figure legends

Figure 1 Flow diagram of the literature search 40 studies were finally included,of which 31 studies were conducted meta-analysis respectively.

Figure 2 The proportion of nosocomial infections among confirm cases of COVID-19, SARS and MERS

Figure 1 Proportions of health care workers among confirmed cases of COVID-19, SARS and MERS

Figure 4 Proportions of nosocomial infections excluding health care workers among confirm cases of COVID-19, SARS and MERS

Figure 5 Proportion of doctors among hospital staff with COVID-19, SARS and MERS

Figure 6 Proportion of nurses among hospital staff with COVID-19, SARS and MERS

Figure 7 Proportion of staff other than doctors or nurses among hospital staff with COVID-19, SARS and MERS

Figure 8 Proportion of health care staff with SARS who did not take protective measures.

